# Using disease symptomatology to guide treatment in patients with central centrifugal cicatricial alopecia: introduction of C-CAT scoring tool

**DOI:** 10.1101/2024.09.22.24314172

**Authors:** Aasheen Qadri, Elizabeth Will, Crystal Aguh

**Affiliations:** The Johns Hopkins University School of Medicine, Department of Dermatology, Baltimore, Maryland

**Keywords:** central centrifugal cicatricial alopecia, alopecia, scarring alopecia, hair loss

## Abstract

**Background:** Central centrifugal cicatricial alopecia (CCCA) is a progressive scarring alopecia. No disease activity scale exists, making assessment of therapeutic intervention difficult.

**Objective:** This study introduces the CCCA Clinical Assessment Tool (C-CAT), a novel scale that quantifies symptom severity to facilitate tailored treatment and track disease progression. Methods: A retrospective review of patients with CCCA were assessed on degree of pruritus, erythema, pain, disease progression, and scalp resistance, each scored from 0-2, over the course of a minimum of six months of therapy.

**Results:** Eighty-two patients were included. The average initial C-CAT score was 3.4, consistent with mild to moderate disease activity. After six months of treatment, 88% of patients had improvement in score, with 48% of all patients achieving remission. Intralesional kenalog injections and topical clobetasol were the most commonly used therapies. Patients required an average of 3.6 different treatments to achieve therapeutic response.

**Limitations:** The subjective nature of some C-CAT components may lead to inter-rater variability.

**Conclusion:** The C-CAT provides a structured, quantitative method to assess CCCA severity, which facilitates tracking of disease improvement for the majority of patients with targeted therapy. This tool promotes personalized, symptom-based treatment and supports clinicians and patients in understanding this complex condition.

**CAPSULE SUMMARY:**

- A novel central centrifugal cicatricial alopecia scoring tool demonstrates that established treatments quantitatively improve symptoms in most patients with central centrifugal cicatricial alopecia over 6 months.
- The novel disease activity scale guides clinicians in structuring treatment based on specific symptomatology and aids patients in understanding progress.

## Introduction

Central centrifugal cicatricial alopecia (CCCA) is a progressive scarring alopecia that exhibits a high disease burden in women of African descent.^1^ While disease activity is often highlighted by pruritus, trichodynia, and, in some cases, erythema, the degree of symptoms, particularly erythema, can often be much less obvious to the patient and clinician, precluding application of other disease activity scales such as the Lichen PlanoPilaris Activity Index (LPPAI) and the Frontal Fibrosing Alopecia Severity Index (FFASI).^2,3^ Perifollicular hyperpigmentation is more commonly seen in CCCA, as opposed to the frank erythema observed in these other cicatricial alopecias. Moreover, the extent of scarring in CCCA can far exceed the perceived degree of inflammatory symptoms, rendering it similar to fibroproliferative disorders, in which even low levels of inflammation can lead to excessive, irreversible fibrosis.^4^ For these reasons, identifying active disease, even when minimal, is critically important. Given that aberrant fibrosis is a defining feature of the disease, treatment should be geared at targeting inflammation and or fibrosis, depending on the unique symptomatic profile of each patient. Standard therapy includes the use of topical or intralesional steroids and oral anti-inflammatory medications such as doxycycline.^5^ Given the nuanced symptomatology and potential for poor outcomes without prompt recognition of disease activity and treatment, both clinicians and patients require a tool to quantitively track CCCA.

To address the lack of validated outcomes tracking in CCCA, we introduce the Central centrifugal cicatricial alopecia Clinical Assessment Tool (C-CAT). This novel scale aims to streamline clinical evaluation of disease activity and guide treatment based on disease activity. The C-CAT assigns a quantitative score to each of the five common presenting signs and symptoms of CCCA. The sum of all symptom scores reflects a measurement of disease activity and can be used to guide therapeutic intervention. Here, we demonstrate the application of C-CAT in our CCCA patient population. Using this standardized tool allows providers to incorporate a C-CAT-based treatment ladder specifically tailored to an individual patient’s disease, and additionally encourages patient understanding of disease status.

## Methods

### Study design

This retrospective chart review was approved by the Johns Hopkins Institutional Review Board. Patients with clinically or histologically confirmed CCCA who were seen between January 2023 and February 2024 with a minimum of two visits over the course of six months or longer were included.

### C-CAT Score Calculation

Each of the four predominant symptoms of CCCA is given an individual score (Figure 1). Pain and pruritus are subjective and scored based on patient-reported frequency and/or intensity of symptoms. Erythema and scalp resistance are objective and measured by the provider at the time of visit. Scalp resistance can only be assessed during scalp injections; therefore, patients who do not receive scalp injections during the visit receive “not available (NA)” for their total C-CAT score. All patients receive a score of 2 for disease progression at the initial visit, given that disease burden was sufficient to prompt medical attention, thus indicating a change from baseline. After calculating scores for each subcategory, a sum is derived to characterize the total disease activity score and corresponds to quiescent, mild, moderate, or severe disease. Treatment can be guided based on both individual symptom score and total C-CAT score.

**Figure 1.**
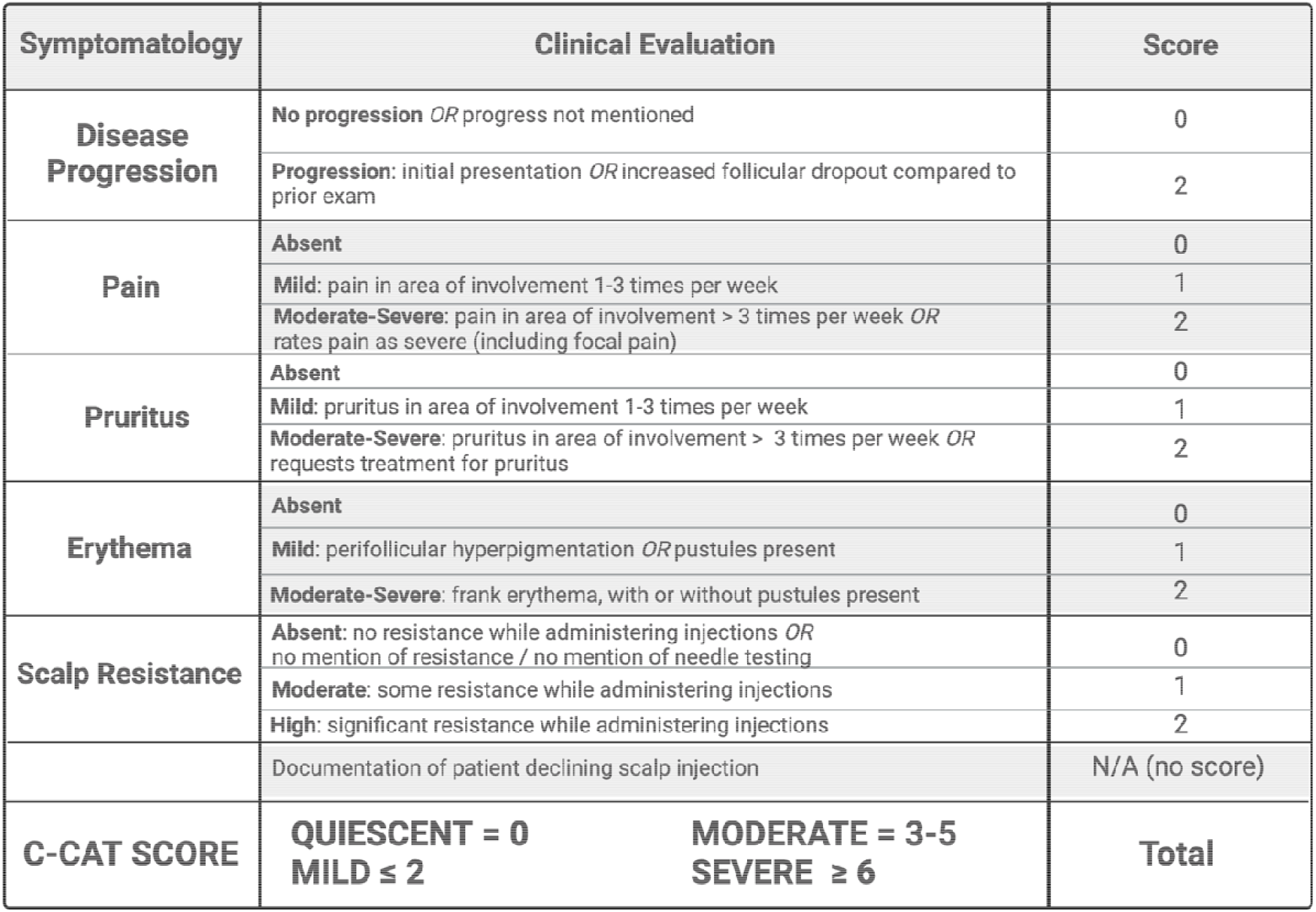
The novel central centrifugal cicatricial alopecia clinical assessment tool (C-CAT). The scale rates clinical metrics from 0 to 2, where 0 represents lack of active symptom, 1 signifies mild symptom, and 2 represents severe symptom. After calculating scores for each subcategory, a sum total is derived to identify the overall disease activity score. The designation of a disease progression score of 2 at the time of first visit impresses upon clinicians the importance of treating appropriately at this initial encounter.

## Results

A total of 82 patients met inclusion criteria. The cohort was 98% (80/82) female, and all patients (82/82) were African-American. Average age at initial presentation was 47.4 years (range 19-83). The most frequent symptom experienced by patients was pruritus 54% (44/82), followed by erythema 39% (32/82), excessive scalp resistance 38% (31/82), and pain 34% (28/82). At initial visit, the average total C-CAT score for all patients was 3.3. As characterized by the C-CAT scale, 37% (30/82) of patients presented with mildly active disease (C-CAT score 1-2), 55% (45/82) with moderately active disease (C-CAT score 3-5), and 8% (7/82) with severely active disease (C-CAT score >6) at the initial visit. Over an average of 195 days, 48% (39/82) of all patients achieved disease remission (score of 0).

Between the initial visit and the most recent visit, 88% (72/82) of patients experienced an improvement in total C-CAT score (“responders”), by an average of 2.7 points. The average C-CAT score was 3.4 at initial visit for responders. On average, patients required 3.6 therapies to achieve response. The most common therapy for responders was ILK injections, received by 96% (69/72) of responders throughout encounters. The next most common therapy among responders was topical clobetasol ointment, prescribed to 68% (49/72) throughout encounters. Compounded topical minoxidil, oral minoxidil, compounded topical metformin, oral metformin, and doxycyline were additional highly utilized therapies (Table 1).

**Table 1.** Responders and non-responders, as described by improvement in C-CAT score, and treatments received.

|  | **Responders (n: 72)** |  |  | **Non-Responders (n: 10)** |  |  |
| --- | --- | --- | --- | --- | --- | --- |
| **Treatment** | **Recived**<br>**during study**<br>**period (%)** | *Received at*<br>*initial visit* | *Received for*<br>*first time at*<br>*follow-up* | **Recived**<br>**during study**<br>**period (%)** | *Received at*<br>*initial visit* | *Received for*<br>*first time at*<br>*follow-up* |
| ILK injections | 96% (69/72) | 71% (49/69) | 29% (20/69) | 90% (9/10) | 56% (5/9) | 44% (4/9) |
| Topical clobetasol<br>ointment | 68% (49/72) | 88% (43/49) | 12% (6/49) | 30% (3/10) | 33% (1/3) | 67% (2/3) |
| Compounded topical<br>minoxidil* | 56% (40/72) | 63% (20/32) | 38% (12/32) | 60% (6/10) | 50% (3/6) | 50% (3/6) |
| Oral minoxidil | 40% (29/72) | 31% (5/16) | 69% (11/16) | 50% (5/10) | 20% (1/5) | 80% (4/5) |
| Compounded topical<br>metformin** | 31% (22/72) | 41% (9/22) | 59% (13/22) | 40% (4/10) | 75% (3/4) | 25% (1/4) |
| Oral metformin | 22% (16/72) | 31% (5/16) | 69% (11/16) | 40% (4/10) | 50% (2/4) | 50% (2/4) |
| Oral doxycycline | 17% (12/72) | 42% (5/12) | 58% (7/12) | 40% (4/10) | 0% (0/4) | 100% (4/4) |

\*Compounded topical includes minoxidil 7%/retinoic acid 0.025%/fluocinolone 0.01% OR minoxidil 8-10%/retinoic acid
0.025%/triamcinolone 0.1% ointment
\*\*topical metformin includes compounded metformin 10-20% in lipoderm cream OR metformin 10%/clobetasol 0.05%

A total of 12% (10/82) of patients experienced either no change in or worsening of their total C-CAT score (“non-responders”). The average C-CAT score at initial visit was 2.6 for non-responders. Disease progression at time of follow-up encounter, marked by increased follicular dropout compared to the prior exam, was identified in 60% (6/10). All but one (9/10) of the non-responders received ILK injections throughout encounters. At time of initial visit, disease was characterized for non-responders as: 50% (5/10) mildly active, 50% (5/10) moderately active, and none (0/0) severely active.

## Discussion

We demonstrate the application of the novel C-CAT disease activity scale to a cohort of CCCA patients, equipping providers with the ability to precisely track symptomatology that is often subtle, provide symptom-based treatment, and facilitate patient understanding of this difficult disease. Our findings support the use of standardized therapy, consisting of anti-inflammatory and anti-fibrotic therapies, as first-line treatments for CCCA.^5^

The majority of responders in this cohort demonstrated score improvement within 6 months, which can support clinicians in setting patient expectations. Improvement was by an average of 2.7 points; this reflects a mix of disease stabilization and symptom improvement. Response to treatment was not predicted by activity level at baseline as all non-responders initially presented with disease that was either mildly or moderately active. This further demonstrates the need for close follow-up of symptoms, even for patients with mildly active disease, who may be at risk for disease progression if symptoms do not become quiescent.

### Treatment algorithm for CCCA can be guided by disease symptoms

A nuanced understanding of pharmacologic interventions can be used to identify the best treatment based on patient symptoms (Figure 2).^6^ The symptoms of pain, pruritus, and erythema experienced by patients are all products of underlying inflammation, suggesting that anti-inflammatories should continue to remain a first line treatment option for this disease. This data suggests that intralesional kenalog injections and topical corticosteroids are indeed effective as first-line therapies in CCCA. Steroids, used for a variety of inflammatory and fibrotic skin diseases such as keloids, result in decreased pro-inflammatory cytokines including TNF-a and IL-1, as well as TGF-β, a pro-inflammatory and pro-fibrotic marker.^7^ For CCCA, mild disease can be treated with ILK injections 2.5-10 mg/cc every 6-12 weeks, and topical steroids such as clobetasol 0.05% ointment 3-5 days per week.^6^

**Figure 2.**
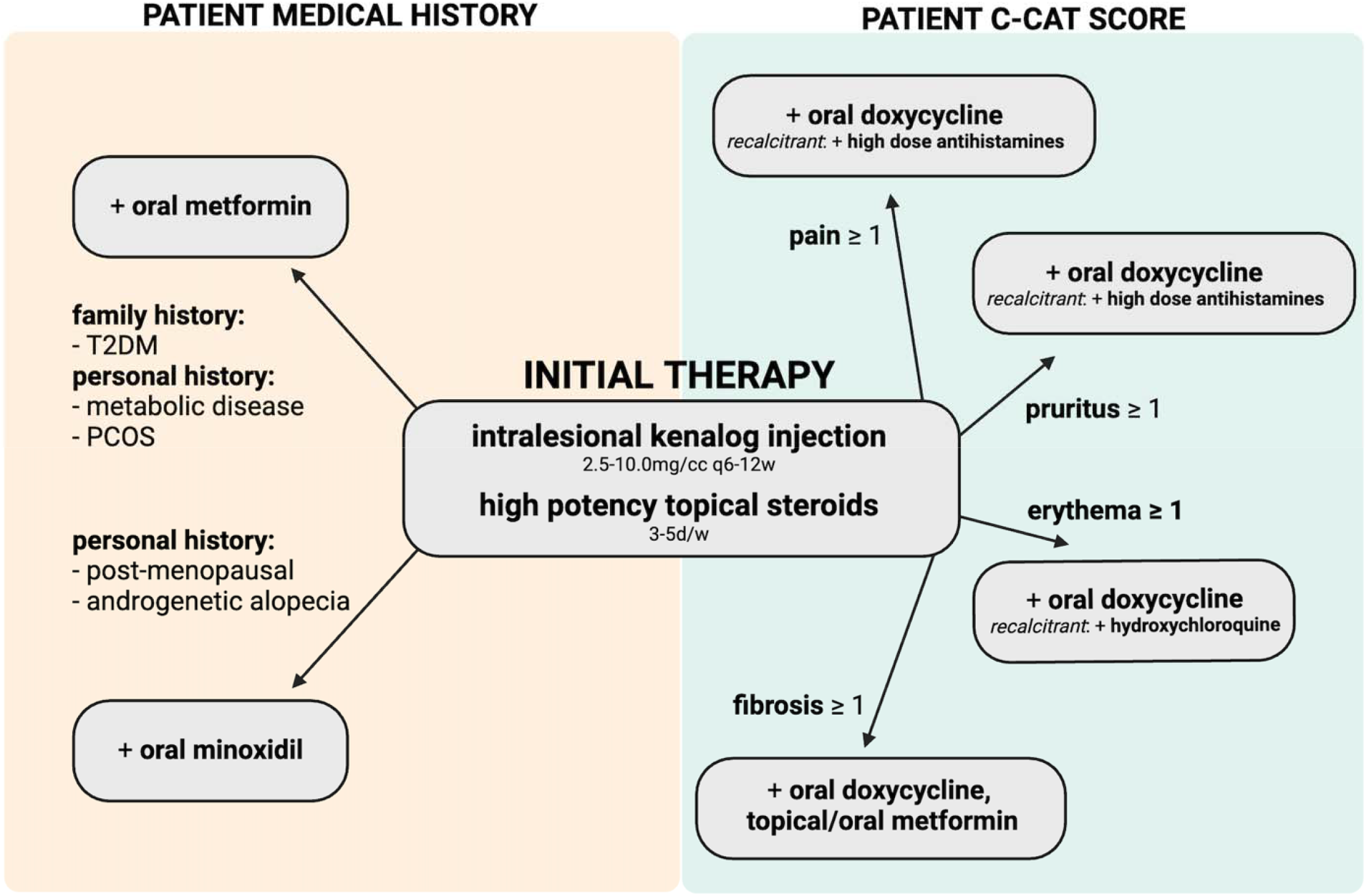
Central centrifugal cicatricial alopecia: Personalization of treatment regimen based on C-CAT score. Initial standard therapy, followed by addition of medication after consideration of factors from the patient’s medical history (yellow) and symptom-specific C-CAT evaluation (green).

Doxycycline is similarly considered a first line treatment for patients with CCCA and is suitable for patients who experience high scores in pain, pruritus, erythema, or scalp resistance. Excess fibrosis and inflammation in CCCA occur through the actions of matrix metalloproteinases (MMPs), endopeptidases that promote extracellular matrix turnover as well as fibroblast activity and stimulation of inflammation.^8^ The upregulation of matrix metalloproteinases in CCCA, specifically MMP1, MMP2, MMP7, and MMP9, suggests a role for doxycycline’s anti-MMP activity.^9^ Doxycycline has been shown to reduce acne scarring via downregulation of MMP-mediated TGF-β production.^10^ Furthermore, tetracyclines have been demonstrated to be effective in decreasing inflammation via reduced PAR-2 signaling.^11^ Gadre et al. found that CCCA patients have higher levels of IL-β1 in their stratum corneum, which induces PAR-2, the hypothesized modulator of cowhage itch. Stimulation of the cowhage itch pathway classically results in burning, tender pruritus, as seen in CCCA.^13^ Therefore, use of tetracyclines to dampen pathway initiation by PAR-2 results in downstream decreased inflammation and itch in pruritic-predominant patients.

Adjunctive, off-label treatment with topical or oral metformin is another therapeutic option for addressing excessive scalp resistance, which can be used as a proxy for fibrosis. Metformin reduces fibrosis primarily by inhibiting the production of TGF-β1, as well as blocking its interaction with its receptor, and activating AMPK to disrupt the phosphorylation and nuclear translocation of Smad2/3, which are crucial for activating fibrogenic genes.^14^ In a study of idiopathic pulmonary fibrosis, AMPK activation by metformin resulted in the apoptosis of myofibroblasts and reversal of established fibrosis.^15^ Moreover, current literature suggests greater clinical benefit with metformin treatment as several risk factors, namely prediabetes and diabetes, are comorbid with CCCA.^16,17^ Although topical metformin has been successful in promoting regrowth of hair in CCCA patients, patients with evidence of insulin resistance may benefit from systemic metformin treatment. Patients with CCCA should be routinely tested for pre-diabetes, with hemoglobin A1C greater than 5.7 or HOMA-IR greater than 2 indicative of insulin resistance. Populations that we propose for systemic treatment include those with lab studies indicative of pre-diabetes, a family history of Type II diabetes, signs of metabolic disease, or a known history of polycystic ovary syndrome (PCOS), in addition to patients with elevated fibrosis scores.

Oral minoxidil can be utilized in patients with concurrent androgenetic alopecia (AGA), which is most common in those patients who are peri-or post-menopausal. CCCA patients with comorbid AGA present with a higher degree of disease burden due to the combined alopecia. Thus, it is imperative to address AGA aggressively, preferably with systemic treatment such as oral minoxidil therapy, which has been proven to be efficacious in addressing decreased hair density as caused by AGA.^18^ Furthermore, it has been suggested that minoxidil may also have some anti-fibrotic effect through reduced activity of the TGF-β/Smad2/3 caspase signaling pathway and inhibition of the lysyl hydroxylase pathway, resulting in decreased collagen formation and deposition.^19^

Other adjunct treatments can be considered depending on each patient’s need. For intractable pruritus, high-dose second-generation antihistamines such as cetirizine can be employed as antihistamines block the pro-inflammatory activity of H1 histamines and reduce the resulting pruritus.^20^ However, only one patient in our cohort was treated cetirizine, as majority reported sufficient relief of their pruritus with steroids and/or doxycycline. For recalcitrant erythema, hydroxychloroquine can be considered as it has been demonstrated to decrease disease activity in other more clinically inflammatory cicatricial alopecias.^21^

A limitation of the C-CAT is the nuance inherent to CCCA symptomatology. For clinicians who do not frequently care for CCCA patients, it may take time to establish a consistent understanding of symptoms. For example, as stated above, erythema can be very subtle, manifesting only as perifollicular hyperpigmentation and in these cases, trichoscopy can be particularly helpful. Additionally, fibrosis has the potential for significant inter-rater variability, but becomes easier to define with time, especially when clinicians become specifically attuned to the degree of resistance that is to be expected in both patients with scarring and non-scarring alopecias (for example, alopecia areata) receiving injections.

However, clinicians will find that introduction of a clinical disease activity scale into daily practice has the potential to drastically improve patient clinical experience. A major benefit of the C-CAT is the ability to nurture the provider-patient relationship by quantitatively demonstrating to patients their improvement with treatment, despite lack of hair regrowth. The C-CAT arms clinicians with evidence to support continued treatment when necessary and maintain hope.

In conclusion, the novel C-CAT provides an objective, quantitative measure to evaluate the most common symptoms of CCCA and guide patient-tailored treatment. Improvements in score, often seen within 6 months, can be encouraging for both patients and clinicians, and reinforce that progress is being made despite the lack of visual hair regrowth in this challenging disease. Future directions include randomized clinical trials to further solidify treatment recommendations and identify predictors for response/remission.

## Data Availability

All data produced in the present study are available upon reasonable request to the authors

## ACKNOWLEDGEMENTS

None.

